# Electronic Computer-Based Model of Combined Ventilation Using a New Medical Device

**DOI:** 10.1101/2021.01.17.21249912

**Authors:** Matias Ramos, Roberto Orofino Giambastiani, Diego Riva, María Fernanda Biancolini, Ignacio Lugones

## Abstract

**Introduction:** The increased demand for mechanical ventilation caused by the SARS-CoV-2 pandemic could generate a critical situation where patients may lose access to mechanical ventilators. Combined ventilation, in which two patients are ventilated simultaneously but independently with a single ventilator has been proposed as a life-saving bridge while waiting for new ventilators availability. New devices have emerged to facilitate this task and allow individualization of ventilatory parameters in combined ventilation. In this work we run computer-based electrical simulations of combined ventilation. We introduce an electrical model of a proposed mechanical device which is designed to individualize ventilatory parameters, and tested it under different circumstances.

**Materials and Methods:** With an electronic circuit simulator applet, an electrical model of combined ventilation is created using resistor-capacitor circuits. A device is added to the electrical model which is capable of individualizing the ventilatory parameters of two patients connected to the same ventilator. Through computational simulation, the model is tested in different scenarios with the aim of achieving adequate ventilation of two subjects under different circumstances: 1) two identical subjects; 2) two subjects with the same size but different lung compliance; and 3) two subjects with different sizes and compliances. The goal is to achieve the established charge per unit of size on each capacitor under different levels of end-expiratory voltage (as an end-expiratory pressure analog). Data collected included capacitor charge, voltage, and charge normalized to the weight of the simulated patient.

**Results:** Simulations show that it is possible to provide the proper charge to each capacitor under different circumstances using an array of electrical components as equivalents to a proposed mechanical device for combined ventilation. If the pair of connected capacitors have different capacitances, adjustments must be made to the source voltage and/or the resistance of the device to provide the appropriate charge for each capacitor under initial conditions. In pressure control simulation, increasing the end-expiratory voltage on one capacitor requires increasing the source voltage and the device resistance associated with the other simulated patient. On the other hand, in the volume control simulation, it is only required to intervene in the device resistance.

**Conclusions:** Under simulated conditions, this electrical model allows individualization of combined mechanical ventilation.

## Introduction

Acute respiratory distress syndrome caused by SARS-CoV-2 increased the demand for mechanical ventilation worldwide [1]. An imbalance between supply and demand for ventilators could pose a critical scenario in which it would be necessary to decide which patients have to be assigned to these devices and which ones not [2, 3, 4].

Combined ventilation has been postulated as a strategy to face this problem [5,6]. Although this setup may be simple in principle, the dynamics of two patients connected to the same ventilator may require deep understanding of advanced concepts related to mechanical ventilation. This fact has led some scientific societies to disencourage this configuration. One of the strongest arguments against this arrangement is the inability to personalize ventilatory parameters for each patient [7].

As interest in combination ventilation increased, the need to configure parameters individually for each patient led to the creation of different devices that facilitated the task. DuplicAR⍰ is one of these devices that allows simultaneous and independent ventilation of two subjects with only one ventilator [8]. The goal of this device is to enable mechanical ventilation of two patients with a single ventilator, without cross-contamination, and allowing for independent management of the inspiratory pressure (Pi) and positive-end expiratory pressure (PEEP). In this work we propose an electrical simulation model of combined ventilation and the in silico test of the DuplicAR⍰ device.

## Materials and methods

### Systems to combine ventilation

Although combined ventilation can be implemented with a particular configuration of the inspiratory and expiratory tubes, in the last year, different devices have emerged that facilitate implementation and add setting possibilities to each individual under combined mechanical ventilation. DuplicAR⍰ is a medical device which functions as a complementary adapter to the mechanical ventilator and provides a mechanism to offer adequate and independent pressurization of the system for the two subjects. It consists of a device with two “Y” pieces and regulators embedded in its structure. The inspiratory adapter connects to the inspiratory port of the ventilator and to each subject’s inspiratory line (Figure 1). Each inspiratory line can control the inspiratory pressure through a diameter (i.e. resistance) regulator that allows independent management of the tidal volume (Vt). The expiratory adapter connects to each subject’s expiratory line and to the expiratory port of the ventilator. Each expiratory line has a positive end-expiratory pressure (PEEP) controller for independent management of this variable. Cross-contamination is prevented through one-way valves and microbiological filters in each line of the circuit.

**Figure 1.**
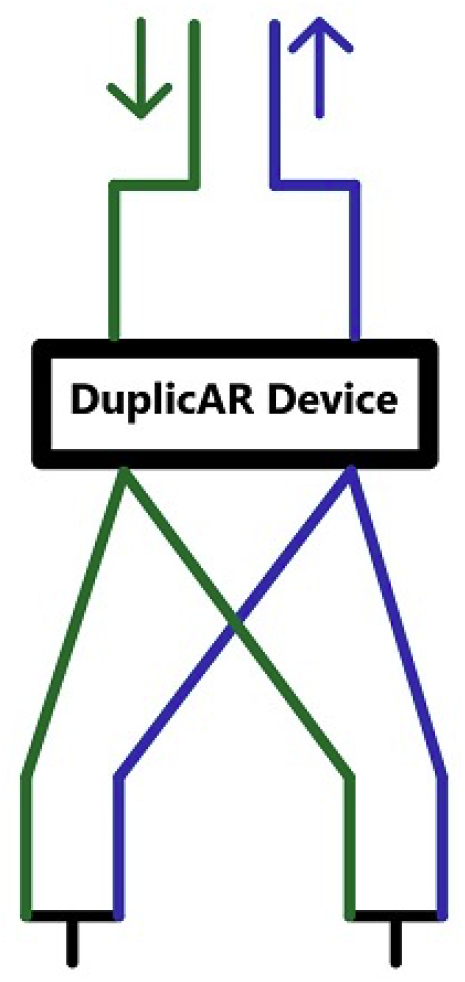
The DuplicAR^®^ device: schematic drawing (green: inspiratory lines; blue: expiratory lines; arrows: ventilator ports).

### Electrical Model Simulation

The code to reproduce the circuits used in the simulations with the corresponding instructions can be found in the supplementary material or in the following link (<u>circuit schematics</u>).

Our electric combined ventilation model is made up of two systems: the “ventilator system” and the “subject system” (Figure 2). The ventilator system includes a constant voltage or current (depending on simulated ventilation mode) and a ground connection. The first is a power source which represents the inspiratory part of the ventilator, while the second represents the expiratory part. The subject system is represented by a series connection of an electrical resistance and a capacitor (RC circuit).

**Figure 2.**
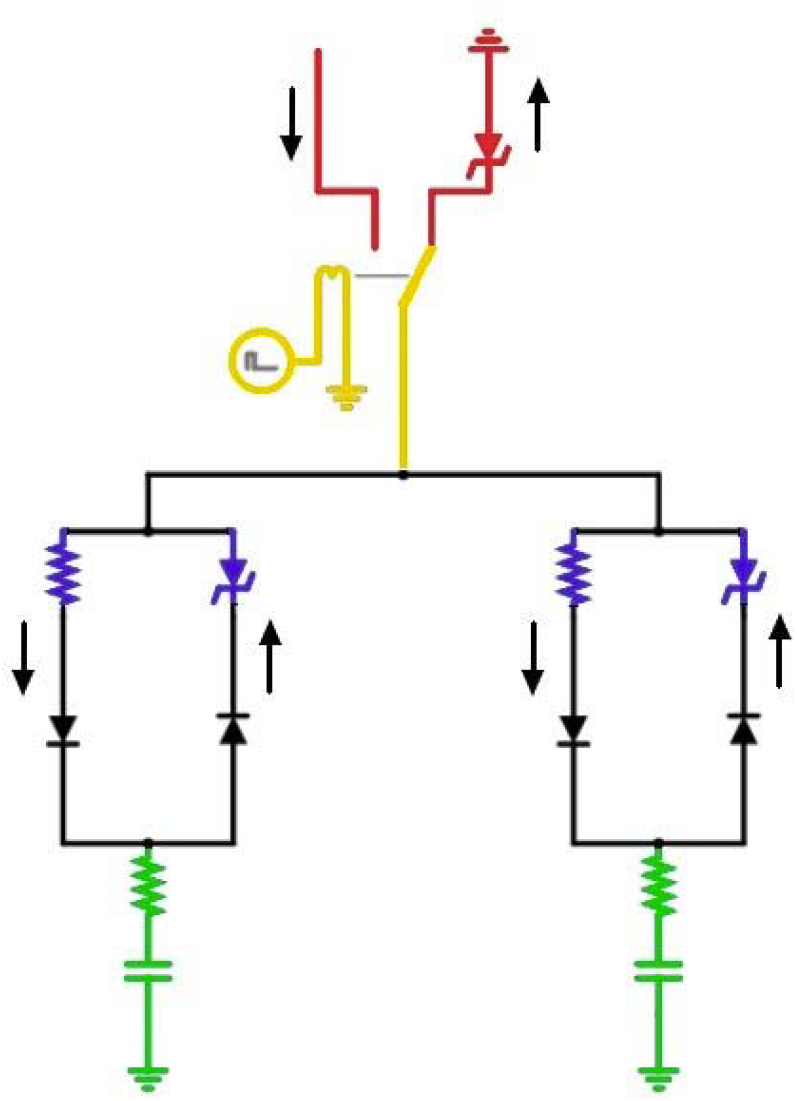
Electrical diagram of combined ventilation: in red, the “ventilator system”; in green, the “subject system”; in yellow, the connection between both systems through the relay; in blue, the DuplicAR^®^ system. The black arrows outside the circuit represent the direction of the current.

The connection between both systems is achieved through a relay. The relay is a device that works as a switch triggered by an external circuit. The current from the external circuit passes through a coil generating a magnetic field that moves the switch of the relay. In this way, the switch moves from the power source to ground in a cyclical manner, governed by the current in its external circuit. By manipulating the current in the external circuit, the behavior of the relay can be controlled. Thus, a ventilatory cycle is simulated by a current pulse in the external circuit that moves the switch (relay) to the power source and then at the end of the pulse, back to ground. The percentage of the cycle that the external circuit source maintains the voltage across the coil (and therefore the switch on “inspiration”) is determined by the “active cycle” of the relay. For example, if the active cycle is set to 33%, an I:E ratio of 1:2 is simulated. The ventilatory rate is represented by the pulse rate. As the “inspiratory port” has a constant voltage source, the system represents the pressure control mode of mechanical ventilation (PCV). When a constant current source is used, the system represents volume control ventilation (VCV). The flows in the inspiratory and expiratory lines are directed by diodes.

To simulate the PEEP, a Zener diode with modifiable breakdown voltage is added prior to grounding the expiratory port. The Zener is a special type of diode that always allows current to flow in one direction, but only allows it in the opposite direction if a certain voltage threshold is exceeded (known as “breakdown voltage”). By setting a zero breakdown voltage, the diode behaves like a simple conductor. Setting a nonzero breakdown value, the diode will “catch” that voltage from “behind” and not allow current to flow to ground. Thus, the Zener diode functions as the equivalent of a PEEP valve.

Figure 3 shows the records obtained during the simulation of PCV (to see the simulation running, video clips can be found in the Supplementary *Material* section).

**Figure 3.**
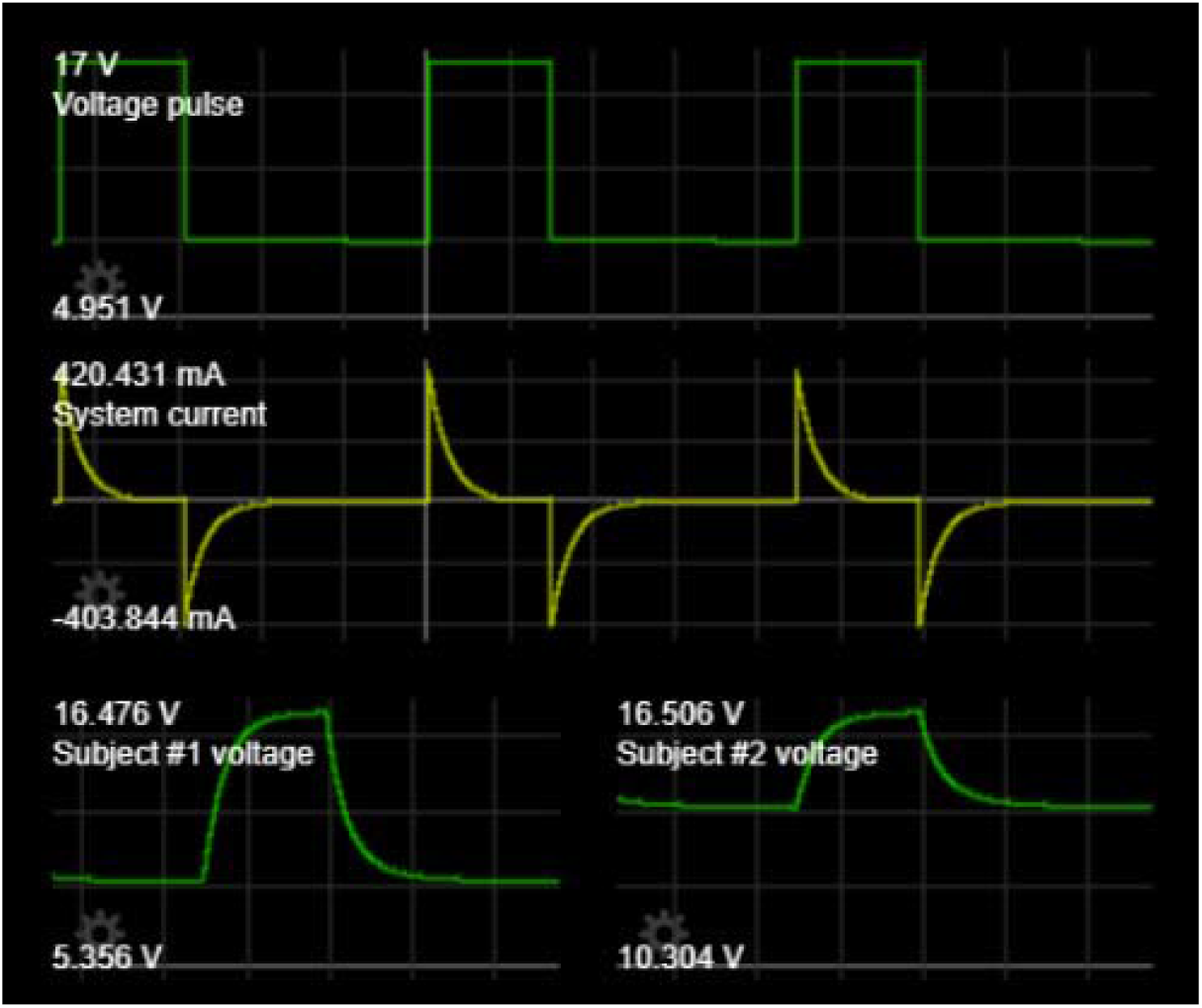
Records in a PCV simulation. All graphs show voltage or current as a function of time. Top: the voltage pulse generated by the “ventilator system”. Medium: the current register in the system. Lower: the capacitor voltage that represents each subject, equivalent to the alveolar pressure in mechanical ventilation.

A theoretical weight is established for each simulated subject. Although weight is not a variable to consider in electric models, in this system we consider weight as the value that determines the amount of charge suitable for the RC circuit. The objective in each simulation is to load that RC circuit with the charge that we consider convenient according to the simulated weight. This normalized charge is chosen at 7 μC kg^-1^, a value that is numerically equal to the 7 ml kg^-1^ that could be used as the tidal volume in real mechanical ventilation. On the other hand, assuming the size of the lung correlates with the size of the individual, we use weight to normalize capacitance to lung size.

Given the “fast” nature of electrical phenomena, the basal cycling frequency of the ventilator system is set at 45 Hz and the active cycle at 33.33%, generating cycles of 22.2 ms (1000ms/45) and “inspiratory times” of 7.39 ms (0.33 ⍰ 22.2). Given the RC circuit characteristics of the simulated subjects, at least 10 expiratory time constants are allowed to take place with this configuration to ensure no charge trapping. This cycling frequency is considered equivalent (in mechanical ventilation) to a respiratory rate of 10 to 12 ventilations per minute in patients with normal resistance and compliance (Table 1) [9].

**Table 1.** Equivalences between the mechanical and electrical models.

| Variable | Real model | Electrical equivalent |
| --- | --- | --- |
| Weight | 50 kg | 50 kg |
| Target volume or charge | 7 ml/kg | 7 $\mu\text{C/kg}$ |
| Compliance or Capacitance | 30 ml/cmH<sub>2</sub>O | 30 $\mu\text{F}$ |
| Resistance | 14 cmH<sub>2</sub>O / l/s | 40 $\Omega$ |
| Cycle frequency | 10 / minute | 45 Hz |
| Inspiration | 2 s | 7.39 ms |
| Expiration | 4 s | 14.81 ms |
| I:E Ratio | 1:2 | 1:2 |
| Time constant ($\tau$) | 0.42 s | 1.2 ms |
| $\tau$ allowed in expiration | 9.5 | 12.3 |

Combined ventilation model involves placing two “patient systems” (ie. two RC circuits) in parallel with a single power source.

The DuplicAR⍰ system is simulated with a variable resistor in the inspiratory line and a Zener diode in the expiratory line of each subject (Figure 2, in blue). By adjusting the electrical resistance of the DuplicAR⍰ system, we control the voltage received by that “patient system”; and adjusting the Zener breakdown voltage of the DuplicAR⍰, we control the end-expiratory voltage (EEV, equivalent to end-expiratory pressure).

The electrical simulations are performed in Falstad Circuit Simulator 2.2.13js, a Java-based electronic simulator [10]. Equivalences between models are shown in Table 1.

### Experimental Setting

Three patient models are simulated, each with particular characteristics of weight and compliance (Table 2).

**Table 2.** Types of simulated subjects.

| Subject / RC circuit | Weight | Resistance | Capacitance | Capacitance/Kg |
| --- | --- | --- | --- | --- |
| A | 50 kg | 40 $\Omega$ | 30 $\mu\text{F}$ | 0.6 $\mu\text{F/kg}$ |
| B | 50 kg | 40 $\Omega$ | 20 $\mu\text{F}$ | 0.4 $\mu\text{F/Kg}$ |
| C | 28 kg | 40 $\Omega$ | 10 $\mu\text{F}$ | 0.35 $\mu\text{F/Kg}$ |

The simulations are run in three different stages, which are recreated by connecting two RC circuits to the same ventilator system and tested under PCV and VCV modes:

*Stage 1*: Two identical RC circuits (A and A), representing identical subjects.

*Stage 2*: Two RC circuits (A and B), representing subjects of the same size but different compliances.

*Stage 3*: Two RC circuits (A and C), representing subjects of different size and compliances.

The initial ventilator setting is configured as follows: 45 Hz of cycling frequency, an I:E ratio of 1:2 and a Zener breakdown voltage of 5 V. In PCV, the inspiratory voltage is configured to ensure charging with 7 μC kg^-1^ each RC circuit. In VCV an inspiratory current is configured in such a way that, given the Ti, the power source delivers the necessary charge to achieve 7 μC kg^-1^ in each RC circuit.

Once ventilation is established, successive 5 V increments are made on the EEV in one RC circuit through manipulation of the Zener diode of the DuplicAR^®^ system. First, voltage is incremented to 10 V of EEV and then up to 15 V, maintaining the initial EEV (5 V) in the contralateral RC circuit and always guaranteeing the same charge of 7 μC kg^-1^ in both. To achieve the objective of the initial configuration and of guaranteeing the 7 μC kg^-1^ for each RC circuit despite the EEV increments, the settings of the DuplicAR^®^ system and/or in the ventilator are adjusted. The data are tabulated and recorded in a spreadsheet for this purpose.

The three stages are summarized in Table 3.

**Table 3.** Summary of the 3 stages, with the characteristics of each subject, initial setting of the ventilator and target charge for each subject.

|  | Stage 1 |  | Stage 2 |  | Stage 3 |  |
| --- | --- | --- | --- | --- | --- | --- |
|  | A | A | A | B | A | C |
| Size (kg) | 50 |  | 50 |  | 50 | 28 |
| Capacitance (μF) | 30 |  | 30 | 20 | 30 | 10 |
| Resistance (Ω) | 40 |  | 40 |  | 40 |  |
| Capacitance/size (μF/kg) | 0.6 |  | 0.6 | 0.4 | 0.6 | 0.35 |
| Initial settings |  |  |  |  |  |  |
| Inspiratory voltage - PCV (V) | 17.5 |  | 23.3 |  | 25.4 |  |
| Inspiratory current - VCV (mA) | 94.5 |  | 94.5 |  | 73.7 |  |
| I:E | 1:2 |  | 1:2 |  | 1:2 |  |
| Cycle frequency (Hz) | 45 |  | 45 |  | 45 |  |
| Target load/size (μC/kg) | 7 |  | 7 |  | 7 |  |
| Target charge (μC) | 350 |  | 350 |  | 350 | 196 |
| Initial EEV (V) | 5 |  | 5 |  | 5 |  |

## Results

### Stage 1

Data is shown in Table 4.

**Table 4.** Data from Stage 1 simulation.

| Stage 1 |  | Initial setting |  | Initial +5 EEV |  | Initial +10 EEV |  |
| --- | --- | --- | --- | --- | --- | --- | --- |
| Subject / RC circuit |  | A | A | A | A | A | A |
| PCV | Inspiratory Voltage (V) | 17.5 |  | 22.5 |  | 27.5 |  |
|  | Vmax (V) | 17.0 | 17.0 | 17.1 | 22.0 | 17.0 | 27.0 |
|  | Vmin (V) | 5.4 | 5.4 | 5.4 | 10.3 | 5.4 | 15.3 |
| | $\Delta V$ (V) | 11.6 | 11.6 | 11.7 | 11.7 | 11.7 | 11.7 |
| | Charge ($\mu C$) | 348.5 | 348.5 | 350.9 | 349.8 | 350.2 | 349.8 |
| | Charge/Size ($\mu C/kg$) | 7.0 | 7.0 | 7.0 | 7.0 | 7.0 | 7.0 |
| | R DuplicAR ($\Omega$) | 0.0 | 0.0 | 160.0 | 0.0 | 275.0 | 0.0 |
|  | EEV (V) | 5.0 | 5.0 | 5.0 | 10.0 | 5.0 | 15.0 |
| VCV | Inspiratory current (mA) | 94.5 |  | 94.5 |  | 94.5 |  |
| | Total charge delivered ($\mu C$) | 700.0 | | 700.0 | | 700.0 | |
|  | Vmax (V) | 17.1 | 17.1 | 17.1 | 22.1 | 17.1 | 27.1 |
|  | Vmin (V) | 5.5 | 5.5 | 5.5 | 10.4 | 5.5 | 15.4 |
| | $\Delta V$ (V) | 11.7 | 11.7 | 11.7 | 11.7 | 11.7 | 11.7 |
| | Charge ($\mu C$) | 350.0 | 350.0 | 349.9 | 350.1 | 349.8 | 350.2 |
| | Charge/Size ($\mu C/kg$) | 7.0 | 7.0 | 7.0 | 7.0 | 7.0 | 7.0 |
| | R DuplicAR ($\Omega$) | 0.0 | 0.0 | 105.0 | 0.0 | 211.0 | 0.0 |
|  | EEV (V) | 5.0 | 5.0 | 5.0 | 10.0 | 5.0 | 15.0 |

It is first evidenced that, without manipulation of the DuplicAR^®^ device, the voltage difference in each capacitor is the same (11.64 ± 0.03 V), and therefore its charge is also the same (349.28 ± 0.85 μC). Charge on each capacitor is calculated according to.

In both ventilatory modes, PCV and VCV, it is possible to modify the EEV of one RC unit (ie. the subject) without compromising its driving voltage or modifying the parameters of the other (Figures 4 and 5). The *driving voltage* is the difference between the maximum and minimum voltage of the capacitor during a cycle, equivalent to driving pressure in mechanical ventilation. To increase the EEV of a RC unit, the Zener breakdown voltage corresponding to that unit (on DuplicAR^®^) must be increased.

**Figure 4.**
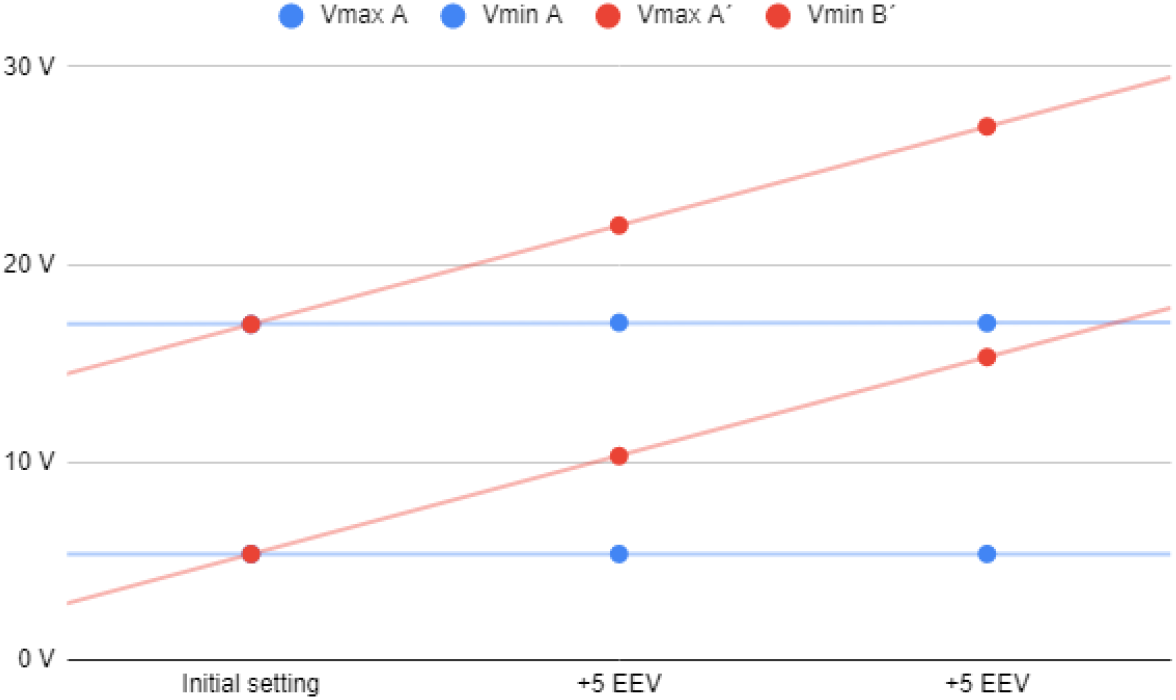
Stage 1 simulation, voltages registers under PCV mode.

**Figure 5.**
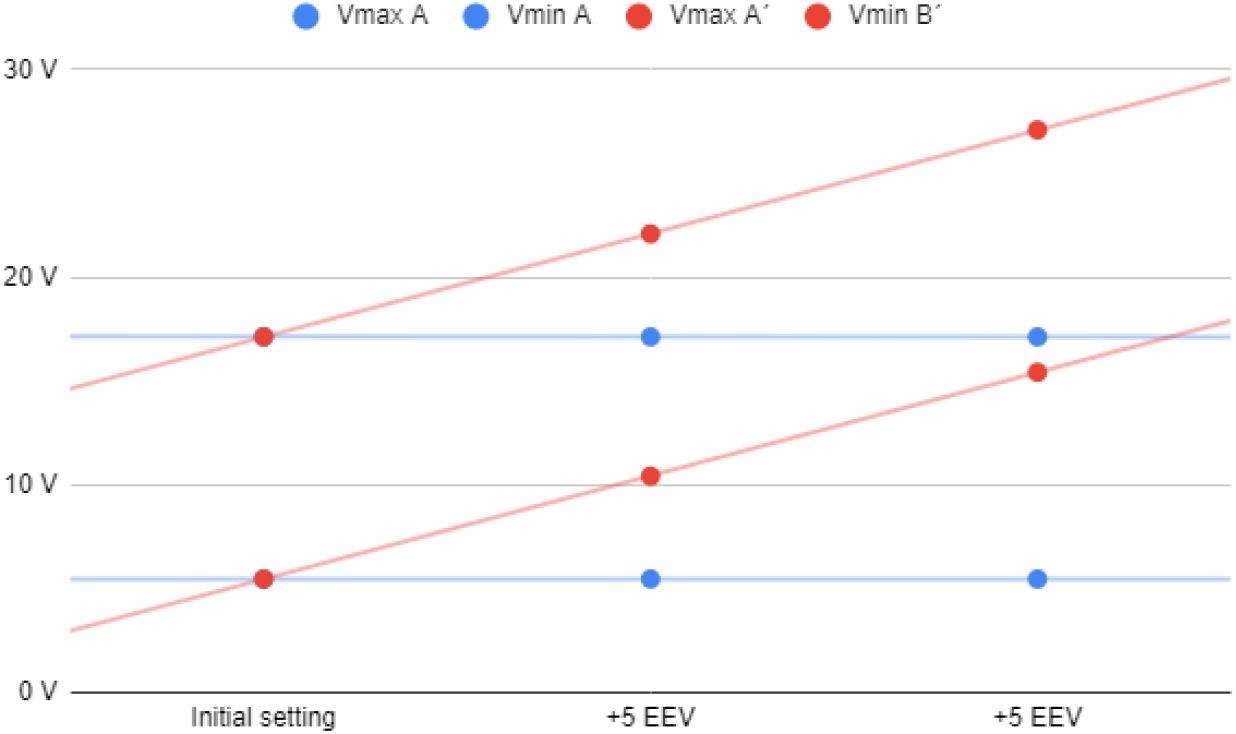
Stage 1 simulation, voltages registers under VCV mode.

With each increase in EEV, the driving voltage is preserved. This is accomplished in PCV mode by two maneuvers: first, increasing the inspiratory voltage of the power source in the same amount as the Zener breakdown voltage; second, restricting the voltage to the other RC circuit by regulating the inspiratory resistance in the DuplicAR^®^ device (Figure 6).

**Figure 6.**
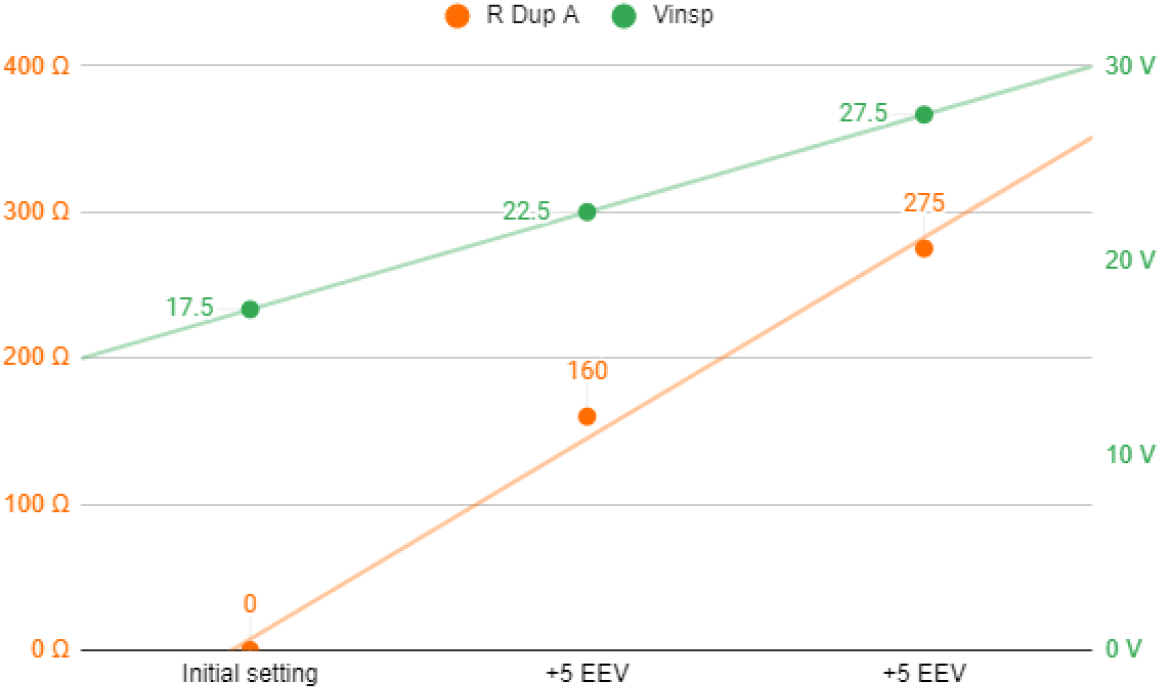
Stage 1 simulation, PCV mode. In green, power source voltage; in orange DuplicAR^®^ resistance of one RC circuit.

In VCV mode, as the EEV of one RC unit increases, it is necessary to adjust the inspiratory resistance of the DuplicAR^®^ device in the other (Figure 7). This is mandatory since, otherwise, no charge would enter the circuit to which the EVV was increased until the breakdown voltage of the Zener diode is reached, causing a disproportionate increase in charge in the second one.

**Figure 7.**
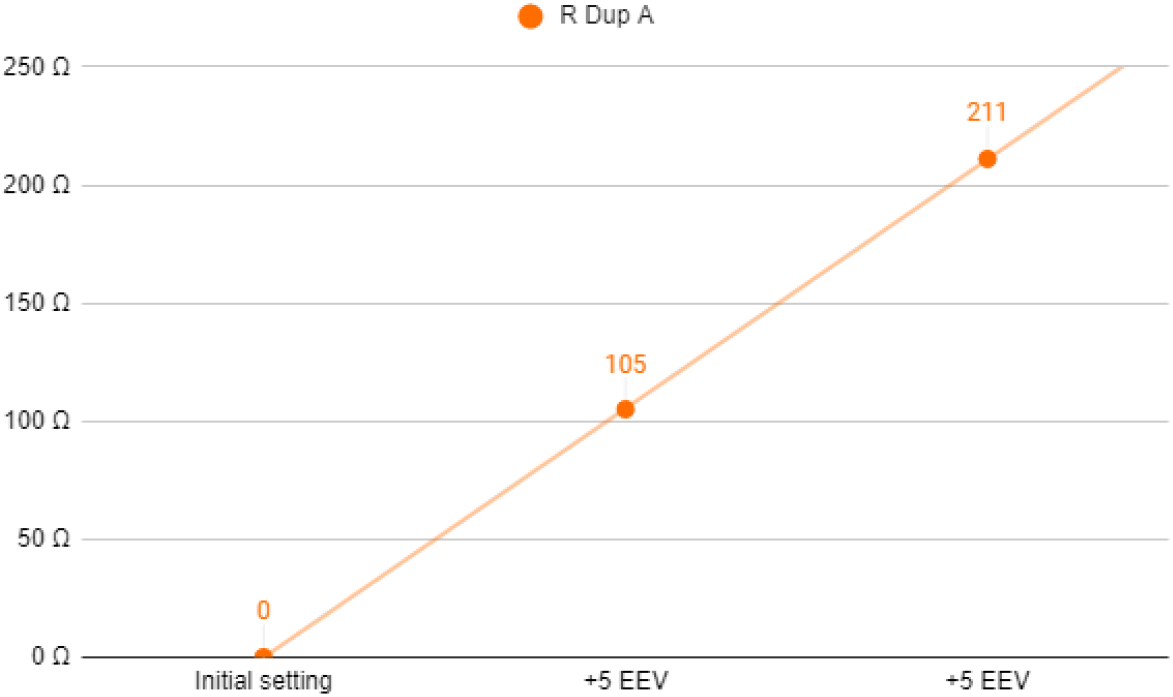
Stage 1 simulation, VCV mode. DuplicAR^®^ resistance of one RC circuit in each step.

### Stage 2

Data is shown in Table 5.

**Table 5.** Data from Stage 2 simulation.

| Stage 2 |  | Initial setting |  | Initial +5 EEV |  | Initial +10 EEV |  |
| --- | --- | --- | --- | --- | --- | --- | --- |
| Subject / RC circuit |  | A | B | A | B | A | B |
| PCV | Inspiratory Voltage (V) | 23.3 |  | 28.3 |  | 33.3 |  |
|  | Vmax (V) | 17.0 | 22.9 | 17.0 | 27.9 | 17.1 | 32.9 |
|  | Vmin (V) | 5.3 | 5.3 | 5.4 | 10.3 | 5.4 | 15.3 |
| | $\Delta V$ (V) | 11.7 | 17.6 | 11.7 | 17.6 | 11.7 | 17.6 |
| | Charge ($\mu C$) | 350.2 | 351.0 | 350.9 | 352.2 | 351.5 | 352.2 |
| | Charge/Size ($\mu C/kg$) | 7.0 | 7.0 | 7.0 | 7.0 | 7.0 | 7.0 |
| | R DuplicAR ($\Omega$) | 180.0 | 0.0 | 292.0 | 0.0 | 400.0 | 0.0 |
|  | EEV (V) | 5.0 | 5.0 | 5.0 | 10.0 | 5.0 | 15.0 |
| VCV | Inspiratory current (mA) | 94.5 |  | 94.5 |  | 94.5 |  |
| | Total charge delivered ($\mu C$) | 700.0 | | 700.0 | | 700.0 | |
|  | Vmax (V) | 17.1 | 23.0 | 17.1 | 27.9 | 17.2 | 32.8 |
|  | Vmin (V) | 5.5 | 5.4 | 5.5 | 10.4 | 5.5 | 15.4 |
| | $\Delta V$ (V) | 11.6 | 17.5 | 11.6 | 17.6 | 11.7 | 17.5 |
| | Charge ($\mu C$) | 349.1 | 350.9 | 349.0 | 351.0 | 350.5 | 349.7 |
| | Charge/Size ($\mu C/kg$) | 7.0 | 7.0 | 7.0 | 7.0 | 7.0 | 7.0 |
| | R DuplicAR ($\Omega$) | 93.0 | 0.0 | 189.0 | 0.0 | 287.0 | 0.0 |
|  | EEV (V) | 5.0 | 5.0 | 5.0 | 10.0 | 5.0 | 15.0 |

Both simulated subjects are the same size, resulting in the same charge requirement (350 μC). Since RC circuit B presents a reduced capacitance, it requires a greater driving voltage than RC circuit A (for circuit A; for B). To achieve these voltage differences in PCV mode, the inspiratory voltage needs to be first configured in the ventilatory system to guarantee the charge to the RC circuit with the lowest capacitance and then to be decreased in the other RC circuit using the inspiratory resistance of the DuplicAR^®^ device (Figure 8, case I).

**Figure 8.**
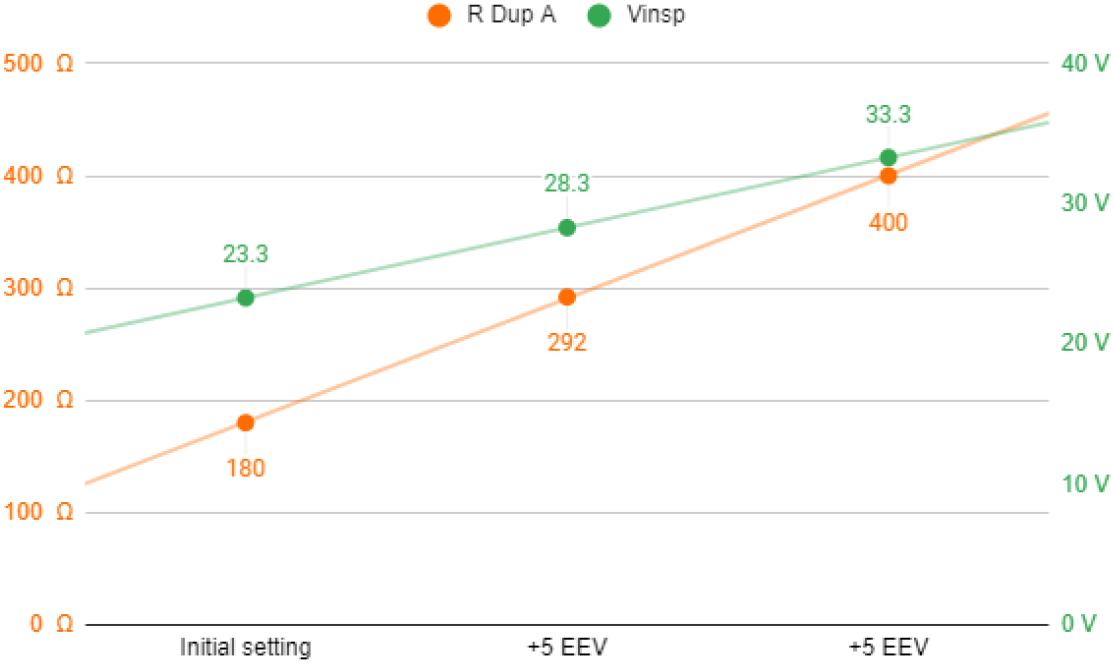
Stage 2 simulation, PCV mode. In green, power source voltage; in orange DuplicAR^®^ resistance of RC circuit A.

In VCV mode, total configured charge is delivered by the ventilatory system to the group, but its distribution results asymmetric regarding differences in RC circuits characteristics. The precise charge distribution for each subject needs to be regulated by the inspiratory resistance regulators of DuplicAR^®^ device in the patient with greater capacitance (Figure 9, case I).

**Figure 9.**
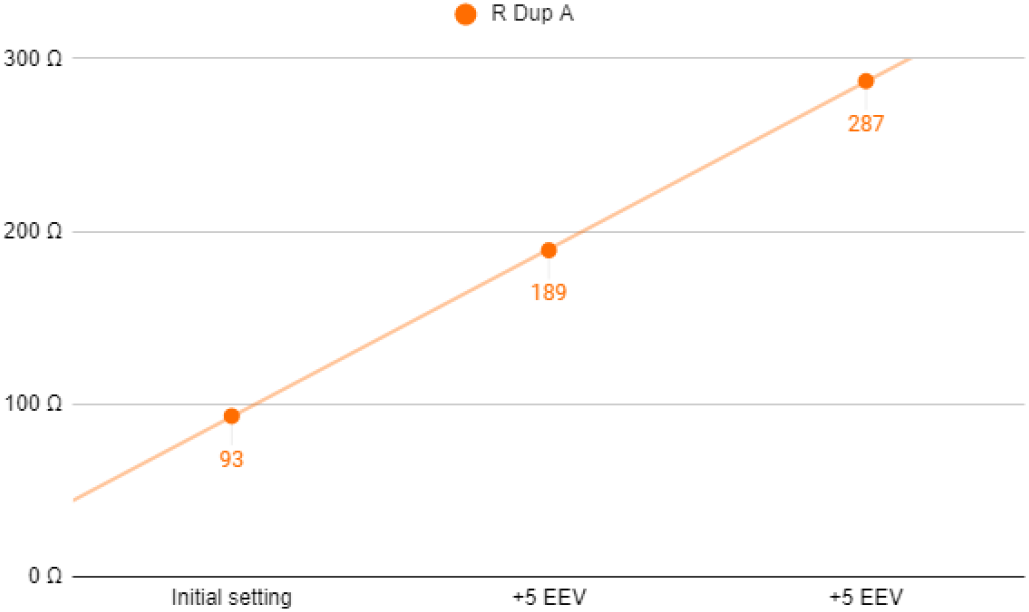
Stage 2 simulation, VCV mode. DuplicAR resistance of RC circuit A in each step.

In this way, with a single ventilator configuration, different driving voltages are established. In both ventilatory modes (PCV and VCV) it is possible to modify the EEV of a RC circuit without compromising its driving voltage or modifying the other circuit’s parameters (Figures 10 and 11).

**Figure 10.**
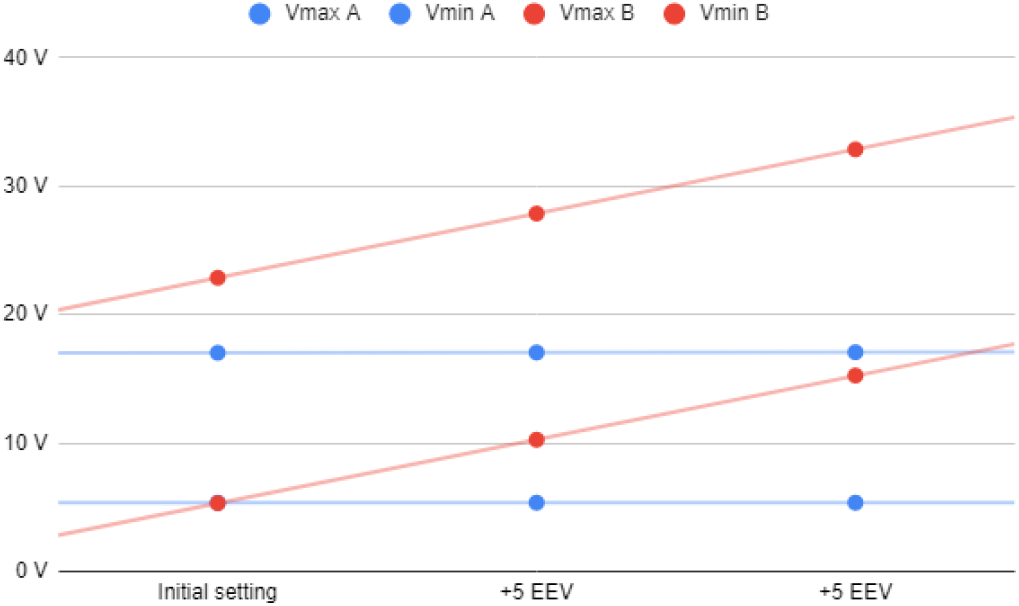
Stage 2 simulation, voltages registers under PCV mode.

**Figure 11.**
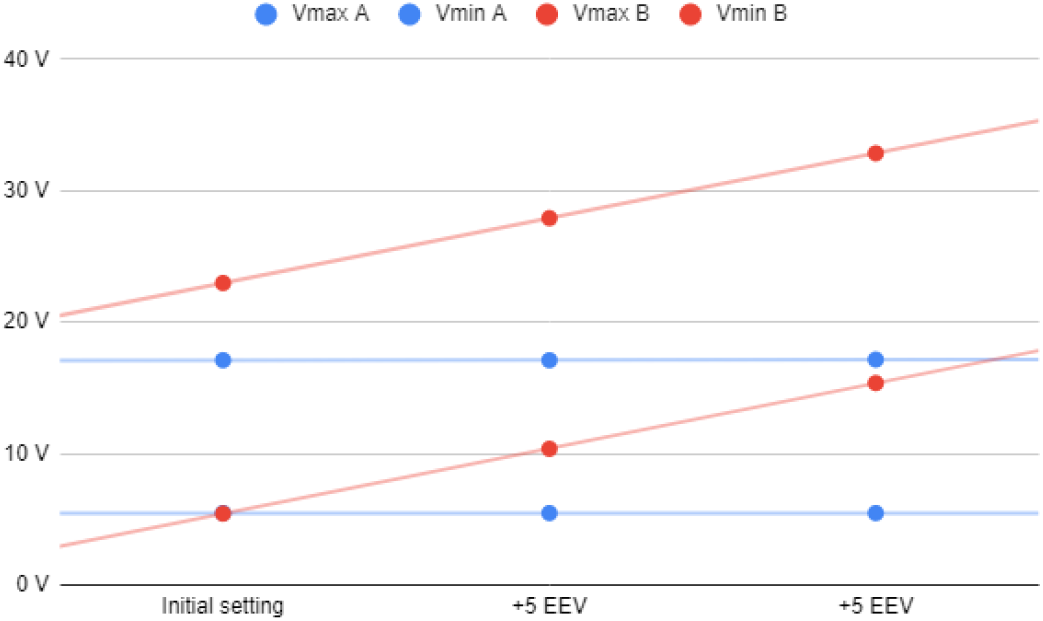
Stage 2 simulation, voltages registers under VCV mode.

With each increase in EEV, the driving voltage is preserved. In PCV mode, this is accomplished by increasing the inspiratory voltage of the ventilator system to maintain the driving voltage of the RC circuit with an increased EEV. Accurate driving voltage in RC circuit A is modulated by the inspiratory resistance of the DuplicAR^®^ device, preventing excessive voltages. In VCV mode the total charge is set. Since EEV of RC circuit B increases, the inspiratory resistance of RC circuit A needs to be adjusted in order to achieve an adequate charge distribution.

### Stage 3

Data is shown in Table 6.

**Table 6.** Data from Stage 3 simulation.

| Stage 3 |  | Initial setting |  | Initial +5 EEV |  | Initial +10 EEV |  |
| --- | --- | --- | --- | --- | --- | --- | --- |
| Subject / RC circuit |  | A | C | A | C | A | C |
| PCV | Inspiratory Voltage (V) | 25.4 |  | 30.4 |  | 35.4 |  |
|  | Vmax (V) | 17.1 | 25.1 | 17.0 | 30.1 | 17.0 | 35.1 |
|  | Vmin (V) | 5.3 | 5.3 | 5.3 | 10.2 | 5.3 | 15.2 |
| | $\Delta V$ (V) | 11.7 | 19.8 | 11.7 | 19.9 | 11.7 | 19.9 |
| | Charge ($\mu C$) | 352.1 | 197.9 | 350.6 | 198.8 | 350.9 | 198.8 |
| | Charge/Size ($\mu C/kg$) | 7.0 | 7.1 | 7.0 | 7.1 | 7.0 | 7.1 |
| | R DuplicAR ($\Omega$) | 226.0 | 0.0 | 339.0 | 0.0 | 447.0 | 0.0 |
|  | EEV (V) | 5.0 | 5.0 | 5.0 | 10.0 | 5.0 | 15.0 |
| VCV | Inspiratory current (mA) | 73.7 |  | 73.7 |  | 73.7 |  |
| | Total charge delivered ($\mu C$) | 546.0 | | 546.0 | | 546.0 | |
|  | Vmax (V) | 17.1 | 25.0 | 17.1 | 29.8 | 17.1 | 34.8 |
|  | Vmin (V) | 5.5 | 5.4 | 5.5 | 10.3 | 5.5 | 15.3 |
| | $\Delta V$ (V) | 11.7 | 19.6 | 11.7 | 19.6 | 11.7 | 19.5 |
| | Charge ($\mu C$) | 349.7 | 196.4 | 350.1 | 195.6 | 350.3 | 195.4 |
| | Charge/Size ($\mu C/kg$) | 7.0 | 7.0 | 7.0 | 7.0 | 7.0 | 7.0 |
| | R DuplicAR ($\Omega$) | 116.0 | 0.0 | 207.0 | 0.0 | 304.0 | 0.0 |
|  | EEV (V) | 5.0 | 5.0 | 5.0 | 10.0 | 5.0 | 15.0 |

It is important to highlight the disparity of voltage differences needed to ensure the target charge of each RC circuit. Capacitor A requires a driving voltage of 11.7 V to reach 350 μC () and capacitor C requires 19.6 V to achieve 196 μC (), reflecting the difference in their *sizes* and capacitances. In both cases the RC units receive the target charge of 7 μC kg^-1^. To achieve these voltage differences in PCV mode, the inspiratory voltage needs to be first configured in the ventilatory system to ensure the charge to the RC circuit with the lowest capacitance and then to be decreased in the other using the inspiratory resistance of the DuplicAR^®^ device (Figure 12, case I).

**Figure 12.**
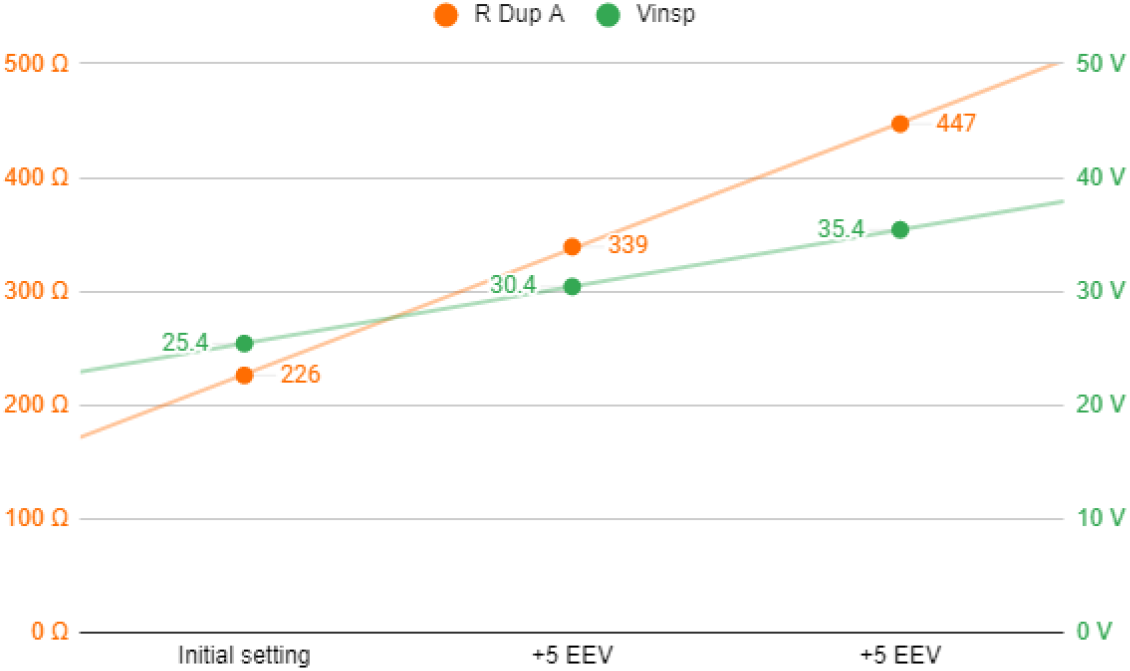
Stage 3 simulation, PCV mode. In green, power source voltage; in orange DuplicAR^®^ resistance of RC circuit A.

In VCV mode, total configured charge is delivered by the ventilatory system to the group, but its distribution results asymmetric regarding differences in RC circuits characteristics. The precise charge distribution for each RC circuit needs to be regulated by the inspiratory resistance controllers of the DuplicAR^®^ device in the unit with greater capacitance (Figure 13, case I).

**Figure 13.**
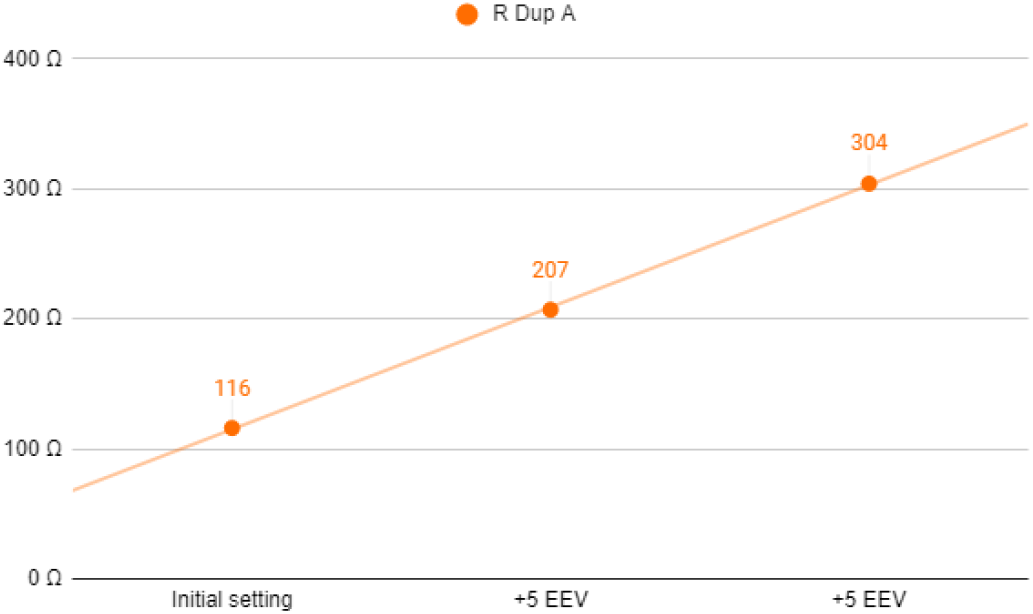
Stage 3 simulation, VCV mode. DuplicAR resistance of RC circuit A in each step.

In this way, with a single ventilator configuration, different driving voltages are established. In both ventilatory modes (PCV and VCV) it is possible to modify the EEV of a RC circuit without compromising its driving voltage or modifying the other circuit’s parameters (Figures 14 and 15).

**Figure 14.**
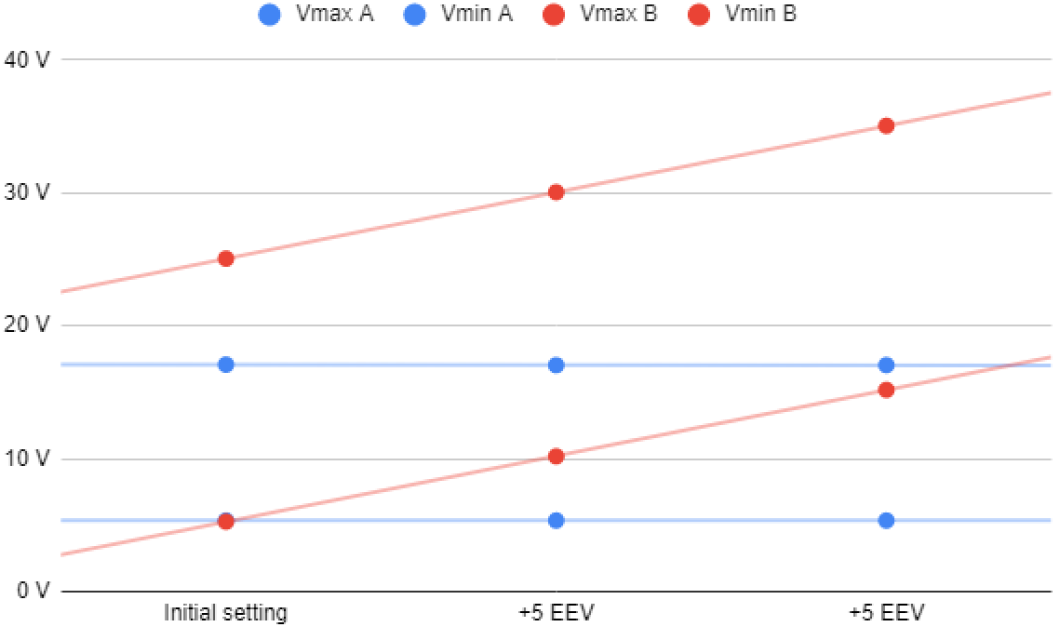
Stage 3 simulation, voltages registers under PCV mode.

**Figure 15.**
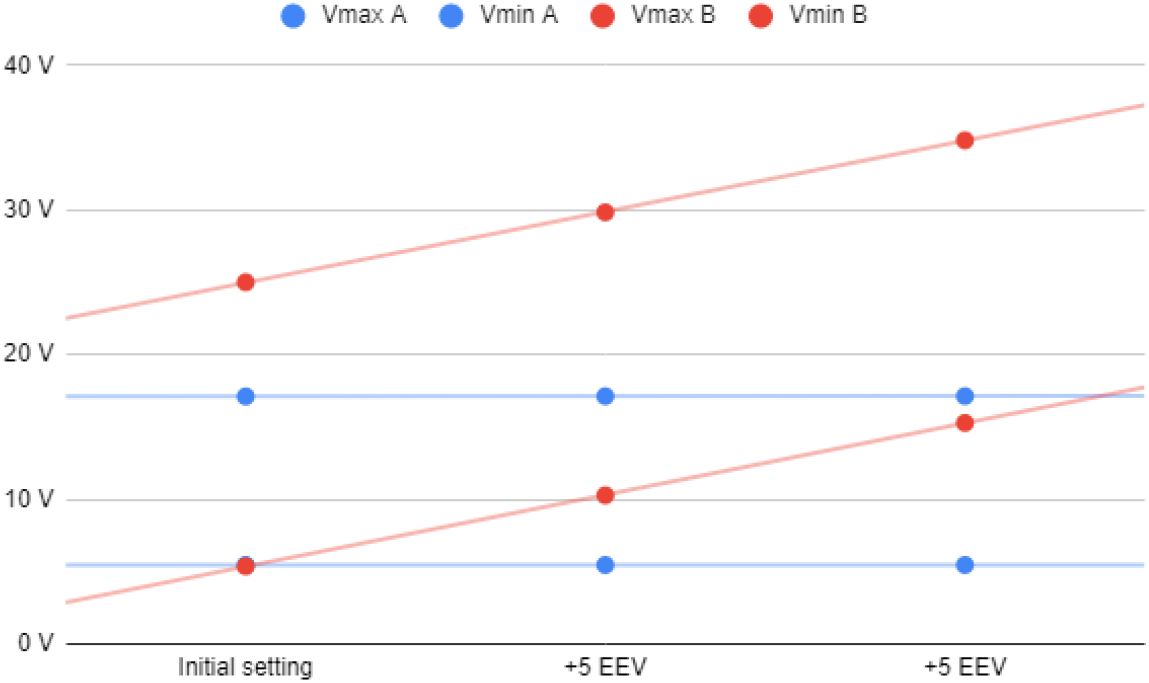
Stage 3 simulation, voltages registers under VCV mode.

With each increase in EEV, the driving voltage is preserved. In PCV mode, this is accomplished by increasing the inspiratory voltage of the ventilator system to maintain the driving voltage of the RC circuit with an increased EEV. Accurate driving voltage in capacitor A is modulated by the inspiratory resistance of the DuplicAR^®^ device, preventing excessive voltages. In VCV mode, total charge is set. Since EEV of capacitor C increases, the inspiratory resistance of RC circuit A needs to be adjusted in order to achieve an adequate charge distribution.

## Discussion

Understanding complex systems can be facilitated by using reductionist models which describe the behavior of variables in simpler ways. With this approach, mechanical ventilation can be represented by an electrical model [11]. By simplifying the respiratory system to an electrical model, we seek to find the relationship between variables of interest (pressure, volume, flow, etc.).

In our simulation system, each variable in mechanical ventilation has an electrical equivalent. The variables pressure, volume, flow, resistance and compliance are considered equivalent to voltage, charge, current, electrical resistance and capacitance, respectively. On the other hand, the components of the ventilatory circuit, such as tubes and valves, are considered equivalent to cables and diodes, respectively.

According to the equation of motion, pressure required to drive gas into the airways and inflate the lungs is caused by the resistive and elastic elements. Lung inflation pressure (P_aw_) can be expressed:

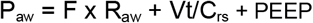

Where F is flow, R_aw_ airway resistance, Vt tidal volume, PEEP positive end-expiratory pressure and C_rs_ compliance of the respiratory system. Using the electrical analog of this equation, the respiratory system can be modeled as an RC circuit, with an electrical resistance connected in series with a capacitor. In these types of circuits, the total voltage (ΔV) should be equal to the sum of voltages on the resistor and capacitor. In this way, the equation that describes the behavior of the system is now:

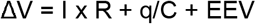

Where ΔV is the potential difference applied to the system, I is the current that flows, R the electrical resistance, q the charge of the capacitor, C the capacitance and EEV is the end-expiratory voltage.

Several studies have addressed the issue of multiple ventilation [12, 13]. In most of these studies, there is no capacity to safely and effectively control the ventilatory parameters for each patient. The only scenario in which adequate combined ventilation is achieved without the need to intervene is in the presence of two identical subjects (same size and compliance) and with the same PEEP. Covid-19 patients’ ventilator requirements can be quite disparate, and also can change over time. When changes in compliance and/or resistance occur, there can be rapid and substantial alteration in the Vt delivered to the other patient.

These types of problems have generated much criticism of combined ventilation and have even discouraged its use. However, in the United States, the Food and Drug Administration has authorized the use of modified ventilator devices and breathing circuits to increase surge capacity (https://www.fda.gov/regulatory-information/search-fda-guidance-documents/enforcement-policy-ventilators-and-accessories-and-other-respiratory-devices-during-coronavirus). This type of modification can be carried out with the DuplicAR device, allowing the individualization of the ventilatory parameters.

Similar modifications to the ventilators have been made and tested with similar results to those shown here. Herrmann and coworkers [15] describe the use of PEEP valves, adjustable constrictions and pressure-relief valves to adapt a ventilator for simultaneous ventilation of two patients. The results of their studies, both in computer simulation and in vitro experimentation, are comparable with those obtained in the present work. The authors do not recommend the “naive” use of combined ventilation as it is particularly dangerous, but they do suggest considering it if devices are used to allow individualization.

Although several works use in silico models to simulate different combined ventilation scenarios, most are based on computational models of mechanically ventilated lungs [16] [17]. Given the analogy between mechanical and electrical systems, we use a simulated electrical model to run different scenarios. The simulations allow future operators an opportunity to understand the systems, manipulate them in real time and push them to the limit without compromising patients. The DuplicAR⍰ system proved to be useful in the ventilation of animals in a previous pilot work (in vivo testing) [8]. Similarly, the device was also evaluated in test lungs (in vitro testing)[18] with results equivalent to those of the present simulation (in silico testing).

The present work shows the performance of the DuplicAR⍰ device in an electronic computer-based model. With this tool it is possible to generate “cleaner” scenarios and even push the device to the limit safely, quickly and with practically no cost.

The results of simulations, transferred to mechanical ventilation, show that it is possible to ventilate two subjects with different sizes and/or compliances and/or PEEP requirements adequately. According to the model, this could be achieved by adjusting the Pi of the ventilator or the PEEP valve of each subject or the inspiratory resistances of the DuplicAR⍰ device. The type and magnitude of the adjustment in each component of the model depend on the ventilatory mode and the characteristics of the subjects.

While these simulations show that the ventilatory goals can be achieved in any of the ventilatory modes, PCV has advantages over VCV. In PCV, the driving pressure can be established and delivered during the inspiratory cycle to both subjects, regardless of their RC characteristics, that is, of their airway resistance and pulmonary compliance. In this mode, there is certainty about the Pi of each subject. As a general limitation of this ventilation mode, it is not possible to know the volume that will be delivered to the system or to each subject. On the other hand, in VCV mode, a total volume to be delivered must be established, that is, the sum of the Vt of both subjects. This volume is not delivered equally to each subject. It is rather distributed among them according to their RC characteristics, and there is no control over the Pi and the Vt.

Although not demonstrated in our simulation, another advantage of PCV mode is that changes in RC characteristics of one subject (i.e. dynamical compliance variability, endotracheal tube obstruction, etc.) do not affect pressurization of the system, and therefore have no consequences on the contralateral subject. On the contrary, in VCV mode, changes in resistance or compliance in one subject directly impact the other subject.

In the model, a similar phenomenon is observed when modifying the EEV in one RC circuit. By increasing the EEV in VCV, the contralateral RC circuit receives a greater charge, since it is necessary that the inspiratory lines reach greater voltage to allow charge to be delivered to the RC circuit with greater EEV. In other words, greater proximal voltage is needed to generate a potential difference that allows charge entry into the RC circuit. This is accomplished at the expense of increasing the charge on the capacitor with no modification of the EEV. In contrast, in PCV, the increase in a RC circuit EEV (without modifying the power source) only affects the unit in which the modification was made. As in the model the changes in EEV do not affect capacitance, the driving voltage is lower and therefore the charge received. This behavior of the electric model to changes in the EEV is consistent with the results of the mechanical model [18].

Additionally, it should be considered that PCV spontaneously compensates for the compliance added to the system by the tubing, which in combined ventilation is expected to contain twice the volume compared to single (conventional) ventilation. This must be manually compensated in VCV mode by adding an extra volume to the total charge delivered by the ventilator. According to the results of the simulations of this study, the most efficient and safest ventilatory mode to use DuplicAR⍰ is PCV.

Although in the theoretical field and in in vitro experimentation, combined ventilation seems promising as a life-saving strategy, we do not yet have large-scale studies that demonstrate its benefit in terms of morbidity and mortality. The decision to use combined ventilation should be made on a case-by-case basis, basing the decision on the best ethical and scientific principles [19]. The medical personnel in charge must obtain the informed consent of the patients and acknowledge the operation of this ventilation strategy in detail to maximize its success.

## Limitations

The present study has limitations. A first observation is that the capacitance for each capacitor in the electrical model is constant. Thus, we assume that ventilation takes place in the region between the lower and upper inflection points of the compliance curve, where it behaves like a linear function. According to this, capacitance is constant, independent of the voltages applied. In clinical practice this would imply that regardless of the pressure applied, the relationship between a volume and a pressure differential would behave as a constant. This assumption may not be real in mechanical ventilation scenarios. In addition, when the simulation takes place at higher voltages, the gas compression that would occur at these simulated pressures was not modeled in the electrical simulation.

As a second limitation, the precise titration of the inspiratory resistance of each patient achieved in the electrical model can hardly be transferred to the real model.

In our electrical model, the ability of the ventilator to “pressurize” the system is not an issue, as the voltage or current source has unlimited capacity. In real life, ventilators may be limited to provide pressurization to larger systems with greater absolute compliances. The electrical model does not consider the greater compliance of a system with larger tubing connections. This would produce a “volume steal” in VCV mode, which was not contemplated in our simulations.

Finally, the objective of the simulated mechanical and electrical models is to represent, in a reductionist way, the interaction between variables in a combined mechanical ventilation scenario. Such models do not take into account other aspects that should be considered in clinical practice, such as hemodynamic status, quality of gas exchange, and the possibility of inadvertent spontaneous ventilation in patients under combined ventilation.

## Conclusions

The electrical computer-based simulations are a safe, pedagogical, and effective tool for understanding combined ventilation. In this model, the DuplicAR^®^ system was shown to be effective in precisely controlling the distribution of charge between capacitors, even in scenarios with different capacitance and with different end-expiratory voltages. These simulations reinforce the fact that it is possible to individualize the ventilatory parameters in two patients connected to a single ventilator.

## Data Availability

The authors confirm that the data supporting the findings of this study are available within the article [and/or] its supplementary materials.

https://drive.google.com/file/d/1lk93BTy4WkNNzaAS4OOxoXoZmTfO7gke/view

## Acknowledgements

Diego Riva acknowledges the support of the National University of La Plata, CONICET and IP-COVID (Fundación Bunge y Born – Agencia I+D+i) in his projects.

## Supplementary Material

Two video clips running the simulations in both PCV and VCV are included as supplementary material. The simulated values are not strictly those used during the experimental protocols. The electrical circuit used and the morphology of the graphs obtained are shown. A web page is also added where the code can be found to reproduce any simulation carried out, with its initial conditions and the diagrams of the circuits used.

## Supplementary Material

- Two videoclips running the simulations
  - https://drive.google.com/file/d/1lk93BTy4WkNNzaAS4OOxoXoZmTfO7gke/view
- Instructions and code to reproduce the simulations
  - https://www.notion.so/mattplate/Electronic-Computer-Based-Model-of-Combined-Ventilation-Schematics-of-Circuits-4fe0faf4b4c7498c814ca18699bc1a1c

